# Amplicon contamination in labs masquerades as COVID19 in surveillance tests

**DOI:** 10.1101/2020.12.08.20244525

**Authors:** Dan Davidi, Susan Fitzgerald, Hannah L. Glaspell, Samantha Jalbert, Stylianos Maheras, Stephanie E. Mattoon, Vanessa M. Britto, Davidson H. Hamer, Giang T. Nguyen, Judy Platt, Cecilia W. Stuopis, Joshua E. Turse, Michael Springer

## Abstract

A cohort of laboratorians with positive SARS-CoV2 test results were uncovered during asymptomatic COVID-19 screening programs at six universities. Follow-up PCR and antibody tests showed that most of these cases were not true COVID-19 infection but instead arose from reverse-transcribed and amplified viral sequences (amplicons) that are generated during research. Environmental testing showed widespread contamination of amplicons in lab spaces including notebooks, keyboards, glasses, and doorknobs. Minimizing instances of amplicon contamination and developing protocols for handling suspected cases are critical to propel research efforts and to avoid diverting university and healthcare resources from patients with COVID-19. Removal of these individuals from the standard testing protocol, per CDC guidelines for positive cases, risks the spread of true infection. We discuss potential prevention and mitigation strategies.

## Introduction

On June 17, a positive SARS-CoV-2 test result was returned at a university associated with this study. The individual had no known exposure to any infected person and the incidence rate in Massachusetts at the time was 0.004%^1^. Further, the individual had no symptoms prior to the test, nor would they develop symptoms in subsequent days. Over the next day, two more employees who worked in the same research laboratory tested positive. They too did not exhibit any symptoms. All three persons worked with the N2 amplicon. As such, the researchers did not believe they were truly SARS-CoV-2 positive and numerous messages started to come to the offices of the university’s Health Services (UHS) and Environmental Health and Safety (EH&S), voicing a concern that this might be a case of amplicon contamination^2^.

In an attempt to determine if this was a true SARS-CoV-2 infection, two additional tests that targeted different regions of the viral genome (N1 and N3) were performed on each individual - all were negative. Although this suggested the individuals might not have had COVID-19, uncertainty regarding the source of the originally detected viral genetic material and the delay between the initial and follow-up RT-qPCR tests led local health authorities to instruct the university to handle the cases as standard infections. The individuals were instructed to isolate, and all their traced contacts were put into quarantine.

Even though these cases were a significant burden on the individuals, the contact tracing teams, and both UHS and EH&S, initially did not pursue further follow-up on these three cases. Over the next several months, a total of 42 similar cases among laboratory workers were identified at universities associated with this study. Each such case triggered a chain of immediate reactions from local authorities^3^, including asking the employees to go into isolation, shutting down lab spaces and putting multiple close contacts from those research communities and the general public in quarantine. Suspiciously, all cases were clustered in individuals that work in or were in close proximity to laboratories that leverage synthetic DNA of the SARS-CoV- 2 virus for various research objectives. All researchers were amplifying DNA which would be detected by their universities SARS-CoV-2 screening program. Given the large number of cases in a relatively small set of schools and the amount of resources being spent on these cases and the disrupted research activity, we considered it critical to determine whether or not these were true COVID-19 cases or workplace amplicon contamination.

## Results

To determine the cause of the RT-qPCR positive tests, we followed-up on 39 out of the 42 individuals by performing different combinations of tests including RT-qPCR with multiple viral targets, whole genome sequencing, and serological tests for the presence of SARS-CoV-2 IgG/M antibodies (**Supporting Table 1**). Repeat testing of the original samples was usually not possible as specimens were not retained by testing facilities after processing due to their large testing volume. Hence, all reported repeat tests were performed on freshly collected samples 1- 3 days following the original positive test results; antibody tests were performed 33 days on average after the initial test result.

All 39 primary positive tests were obtained by RT-qPCR assays designed to detect the N2 and/or N1 viral genes in anterior nares swabs. Broken down by cases, 29 individuals were negative on all follow-up tests. Another 7 individuals had a positive RT-qPCR follow-up test for N2 but were negative for all other tested targets (i.e., N1, N3, E, and/or RdRp). Notably, the negative qPCR result for 25 of the 36 (29+7) cases was obtained as little as 1-3 days after their initial positive test. We also note that those individuals exhibited high Ct values for N2 already in their initial positive test (36.7±1.7; for cases where Ct values were reported), already suggesting that those individuals may not have been true COVID-19 infections. Sealing the case that the vast majority of these cases were amplicon contamination, 18 of the 19 individuals that were negative when tested for SARS-CoV-2 IgG/M antibodies ∼30 days after the initial positive result (**Supporting Table 1**).

It is extremely important to note that not all cases were positive due to amplicon contamination. We identified a total of 4 positive cases in our cohort. One asymptomatic individual did not receive a follow-up qPCR test but was positive for SARS-CoV-2 antibodies ∼two months after the initial positive result. Two of the individuals had repeat tests that were positive on secondary tests with different amplicons, one of the two individuals was symptomatic. The initial Ct value for the asymptomatic COVID-19 infected individual was 31.3 considerably lower than the Ct in each of the amplicon contamination cases. The fourth individual originally was more complicated. This originally asymptomatic individual was tested on September 24 as positive for N2 with a high Ct value (37.4) and negative for N1 in the same test. An antibody test a month later (October 22nd) came back negative, strongly arguing this was a case of amplicon contamination. On November 10 the individual became symptomatic and tested positive for N1 and N2. This individual is therefore one who was initially positive due to amplicon contamination but then contracted COVID-19 afterwards.

The last case highlights the importance of this study in helping to identify cases of amplicon contamination rapidly. In accordance with CDC guidance, testing programs exclude individuals who tested positive for 90 days following the positive result due to persistent RT-qPCR after SARS-CoV-2 infection^4,5^. These individuals, subsequent to amplicon contamination, can contract COVID-19 and be missed because they are not tested. Luckily, this person was asked to return to routine testing once his serological test returned and shortly thereafter he became symptomatic, presented for clinical evaluation, and was tested positive. As part of our mitigation strategy, we argue that all individuals identified as amplicon contamination be immediately returned to asymptomatic testing regimes.

### Evidence for amplicon in lab environments

In light of the widespread evidence for amplicon contamination in these cases, we tested whether we could detect amplicon contamination in the research environments. Environmental swabs were conducted in 11 locations including lab, office and kitchen spaces with a total of ∼90 different sites tested. Amplicons were found in high titers (Ct<30) on a number of sites including centrifuges, pipettes, gel areas, and bench spaces, as well as on other lab equipment including microscopes and incubators (**Supporting Table1**). Importantly, substantial titers were also found on doorknobs, lab notebooks, pens, glasses and computer keyboards with Ct values ranging from 25.8 to 42.6 with a mean of 32.1±4.9.

The presence of amplicons on doorknobs and bench spaces in neighboring labs, in which SARS-CoV-2 work is not conducted, highlight how individuals in adjoining spaces were amplicon positive. The presence of amplicons on personal belongings could help explain two surprising cases. In one case, a researcher extracted large amounts of DNA encoding for the viral N2 protein from bacteria. The same day that individual tested positive, their roommate, who does not work in a research lab, also tested positive. Both cases were verified as amplicon contamination by the lack of IgG/M antibodies against SARS-CoV-2 and follow-up negative RT-qPCR (against the viral amplicon N1, N2, E and RdRp; **Supporting Table1**). A second case proceeded similarly but the researcher was performing RT-LAMP and the negative follow-up assays were for N1 and N3 and independently N1 and N2.

We stress that amplicon contamination is not a public health risk or likely to lead to positives in the general public. Instead, it is specific to research environments that work with the genetic material of the virus. However, the social and economical burden associated with every positive SARS-CoV-2 case is substantial. Minimizing instances of amplicon contamination in universities and developing protocols for handling suspected cases are critical both for the research community and the general public. We therefore discuss potential strategies and testing regimens that should be applied in cases of suspected amplicon contamination.

### Prevention strategies

We believe a large number of the amplicon positive cases are due to laboratory procedures that lead to amplicon contamination. Amplicon contamination is a well-known issue in laboratories, especially clinical labs^2^. Clinical labs implement a number of protocols to largely eliminate amplicon contamination^6^. These include steps such as performing all pipetting in negative pressure hoods, UV light and bleach cleaning of equipment and work areas, avoiding opening tubes after the amplification reaction, and pipetting below the liquid line or in hoods, In addition, individuals can take a number of steps to lower the chance that amplicon contamination registers during surveillance testing such as testing at the beginning of the day before entering the lab, showering and scrubbing hands with soap and water (not alcohol rub, which does not destroy amplicons) prior to sample collection, and wearing a new pair of disposable gloves when handling test kits, especially if using a self-swab approach^7^. A consortium of biosafety committees and EH&S at our universities, in conjunction with local and state public health departments have been working on a shared guidance policy which will include standardized training to minimize contamination and recommendations for regular documentation of cleaning within these spaces.

### Post-positive mitigation strategies

While all amplicon contamination would ideally be eliminated by improved laboratory protocols, it is unlikely that all cases will be eliminated. Hence, it is necessary to be able to empirically separate between COVID-19 viral infection and amplicon contamination. We advise that two follow-up tests be conducted; ideally, follow-up tests should be performed with tests that probe different amplicons. Because during the initial phase of the infection viral titer can be low^5^ and different amplicons and platforms can be more sensitive than others, we do not recommend retesting the initial sample unless the relative efficiency of each test is known. Instead, we advise 24 hours before collecting the follow-up test to help distinguish between low viral titer early in infection and amplicon contamination. This is because viral titer rises significantly after it is initially detectable by qPCR^5,8^. Waiting 24 hours should also help in cases where the source of amplicon contamination is suspected to be whole genome amplification instead of a smaller part of the viral genome. While several other approaches can be useful to determine if there is amplicon contamination, e.g. qPCR amplification with no RT, we do not advise these as a follow-up strategy as someone can simultaneously have SARS-CoV-2 infection and amplicon contamination.

Finally, one key point is that in communities where there is a large amount of biomedical and life science research underway, it is important during the case investigation process to elicit the specific nature of the work when an individual works in a biomedical laboratory. Investigators should ask about an individual’s occupation, whether it involves any research related to SARS-CoV-2 or if their worksite is in close proximity to a lab where such research is conducted, and if so, inquire about laboratory practices. In addition, all cases should be asked if a member of the household is involved in SARS-CoV-2 research in any way, or if they work in a space adjacent to a lab conducting this research. An attempt should be made to understand if there is potential for amplicon contamination that could yield a positive result with the original PCR assay used. Cases who work in these environments or who have household contacts working in these environments should be offered confirmatory testing with an alternate PCR assay that uses different probes, as discussed above. It should be noted that while most of our amplicon contamination occurred with N2 there is no reason to believe that amplicon contamination isn’t occurring with all amplified portions of SARS-CoV-2. In summary, we hope these prevention and mitigation strategies will help to alleviate the burden being caused to research universities and hospitals and allow us to focus resources on true positive cases without impacting research and researcher safety.

## Methods

### RT-qPCR analysis

All RT-qPCR assays were performed by CLIA labs. Several different primer and probe pairs were used to detect the targets listed in **Supporting Table 1**. Reported Ct values are listed in the **Supporting Table 1**; no Ct value threshold was applied for positive test calling.

### Antibody testing

Convalescent Serum IgM/IgG antibody studies were completed on a total of 19 cases. Assays (VITROS Immunodiagnostic Products; Anti-SARS-CoV-2 Reagent Pack assay.) were performed on samples collected on Day 7-75 post the initial positive test results.

### Swab Sampling Procedure

All test sites are listed in **Supporting Table 1**. At each site, an area of 12 in^2^ was sampled with a sterile 3/16-inch-thick medical grade polyurethane swab (Puritan, Guilford; Cat No. 2 5-1607 1PF SC). Swabs were stroked in vertical and diagonal ‘S’ shapes. New pairs of gloves were used for each sample to avoid cross contamination and were placed in a screwcap 1.5 mL tube pre-aliquoted with transport medium (VMA from AHDC Molecular Diagnostics). Samples were labeled and submitted to AHDC Molecular Diagnostics for analysis by qPCR (no RT) using primers for N1, N2, S and Orf1 viral genes.

## Data Availability

all data presented in this paper is available on submission as a supporting excel file

## Acknowledgments

We thank the COVID Testing Laboratory at the Animal Health Diagnostic Center at Cornell University, Ithaca, NY. We also thank the many staff of the affiliated university health services, environmental health and safety units, and research laboratories who responded to these cases and helped to accumulate the data necessary for this manuscript. D.D is an EMBO long□term fellow (ALTF 1146□2018), an HFSP fellow (LT000232/2019□L) and a Rothschild fellow.

